# Simulation of aerosol and droplet spread during upper airway and gastrointestinal endoscopy

**DOI:** 10.1101/2021.04.20.21255743

**Authors:** Johannes Heymer, Florian Dengler, Alexander Krohn, Christina Jaki, Tobias Schilling, Martina Müller-Schilling, Arne Kandulski, Matthias Ott

**Author notes:** <u>Corresponding author:</u> Dr. Matthias Ott, Department of Interdisciplinary Emergency and Intensive Care Medicine, Klinikum Stuttgart, Kriegsbergstr. 60, 70174 Stuttgart, Germany. These authors share authorship (Arne Kandulski, Matthias Ott).

## Abstract

**Objective:** Aerosols and droplets are the main vectors in transmission of highly contagious SARS-Cov-2. Invasive diagnostic procedures like upper airway and gastrointestinal endoscopy have been declared as aerosol generating procedures. Protection of health care workers is crucial in times of COVID-19 pandemic.

**Methods:** We simulated aerosol and droplet spread during upper airway and gastrointestinal endoscopy with and without physico-mechanical barriers using a simulation model.

**Results:** A clear plastic drape as used for central venous access markedly reduced visualized aerosol and droplet spread during endoscopy.

**Conclusion:** A simple and cheap drape has the potential to reduce aerosol and droplet spread during endoscopy. In terms of health care worker protection, this may be important particularly in low- or moderate-income countries.

## Introduction

Since severe acute respiratory syndrome coronavirus 2 (SARS-CoV-2) is highly contagious, health care workers (HCW) are at risk for infection during COVID-19 pandemic[1–3]. The use of personal protection equipment (PPE) reduces infection risk for HCW[1,3]. Therefore, adequate PPE is crucial for reducing morbidity and potential mortality and maintaining healthcare system capacity[1,4,5]. Respiratory aerosols and droplets are the main vectors in SARS-CoV-2 transmission and pathogenesis[6–9]. There is still a matter of debates on aerosol generating medical procedures. Invasive diagnostic procedures like esophagogastroduodenoscopy (EGD) have been declared as aerosol generating procedures (AGP) by different professional societies[9–17]. With that knowledge, interests in infection-prevention for HCW being highly exposed to droplets and aerosols have been increased during the different stages of SARS-CoV-2 pandemic. Besides adequate PPE, contact time and proximity, mechanical barriers are effective strategies to reduce infection risk[9].

In our study, we visualized potential aerosol and droplet spread during upper airway endoscopy with or without physico-mechanical protection.

## Materials and Methods

We used a modified manikin for difficult airway management to simulate, visualize and photo documentation of aerosol and droplet spread during upper airway endoscopy which has been used previously for this purpose during airway management and cardiopulmonary resuscitation[18]. A standard bronchoscope (Olympus BF Type P40, Olympus Europa SE & Co. KG, Hamburg, Germany) was placed into the pharynx of the modified manikin by a professional to simulate the starting position of upper gastrointestinal endoscopy or bronchoscopy, respectively. To simulate the aerosol spread during pharyngeal intubation during upper endoscopy (regardless if upper gastrointestinal or upper airway endoscopy), no further intubation of the trachea-bronchial system was performed. A modified mucosal atomization device (LMA MAD Nasal™, Teleflex Medical, Durham, NC, USA) attached to a syringe was placed in the manikin’s airway for achievement and simulation of aerosol and droplet spread. Ultraviolet light detectable disinfection detergents (Bode visirub® conc., Bode Chemie GmbH, Hamburg, Germany; Schuelke S&M® Optik, Schuelke & Mayr GmbH, Wien, Oesterreich) were spread by this system with manual pressure on the syringe. Aerosol spread was visualized directly using ultraviolet flashlights. Droplet collection was visualized by ultraviolet light on a white impermeable plastic sampler that was attached to the N95 respirator of the investigator. Endoscopy was performed in a standard manner without any precautions and compared to investigations with physico-mechanical protection by a clear plastic drape (Fenestrated drape transparent, B. Braun Melsungen AG, Melsungen, Germany) perforated by a scissor for endoscope passage covering the manikin or the applied oxygen mask respectively. Two cuts on a standard oxygen mask using a scissor facilitated endoscope passage.

### FOTO AVAILABLE ON REQUEST – EXCLUDED BECAUSE OF PREPRINT SERVER POLICY

Fig. 1: Modified manikin for aerosol and droplet simulation (left) and medical professional with PPE primed with impermeable plastic droplet sampler.

### FOTO AVAILABLE ON REQUEST – EXCLUDED BECAUSE OF PREPRINT SERVER POLICY

Fig. 2: Endoscopy starting position using a covering drape (left). The endoscopy passing the clear drape to the manikin’s mouth (right).

### FOTO AVAILABLE ON REQUEST – EXCLUDED BECAUSE OF PREPRINT SERVER POLICY

Fig. 3: Endoscopy using the drape covering the manikin with an applied oxygen mask (left). Perforated drape and oxygen mask by scissor for endoscope passage (right).

## Results

As visualized in figure 4, our model clearly visualized aerosol exposure during standard upper endoscopy. The proximity of the investigator’s face to aerosol and droplets exposure is remarkable.

**Fig. 4:**
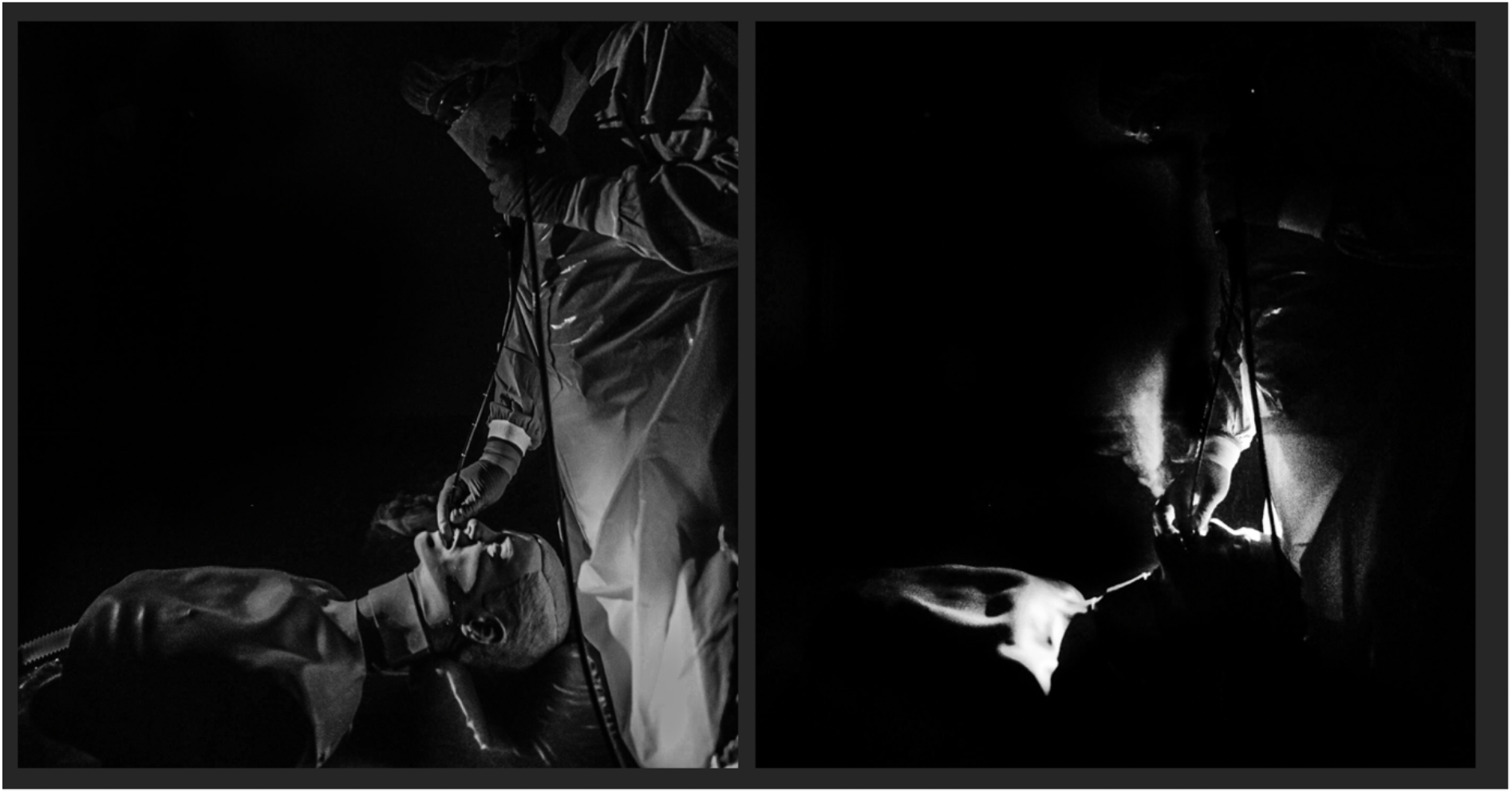
Visualized potential aerosol spread during standard endoscopy.

Having a closer look at the exposure to the HCW (Fig. 5), white spots and dusting on the tracer indicate a notable amount of detectable droplet and aerosol exposure during the procedure.

### FOTO AVAILABLE ON REQUEST – EXCLUDED BECAUSE OF PREPRINT SERVER POLICY

Fig. 5: Droplet and aerosol spread during standard endoscopy and exposure to the HCW. White spots indicate droplets detected by ultraviolet light.

Figure 6 highlights the aerosol spread during endoscopy with a covering clear drape. Aerosol was deflected and could not be detected near the investigator’
ss face.

**Fig. 6:**
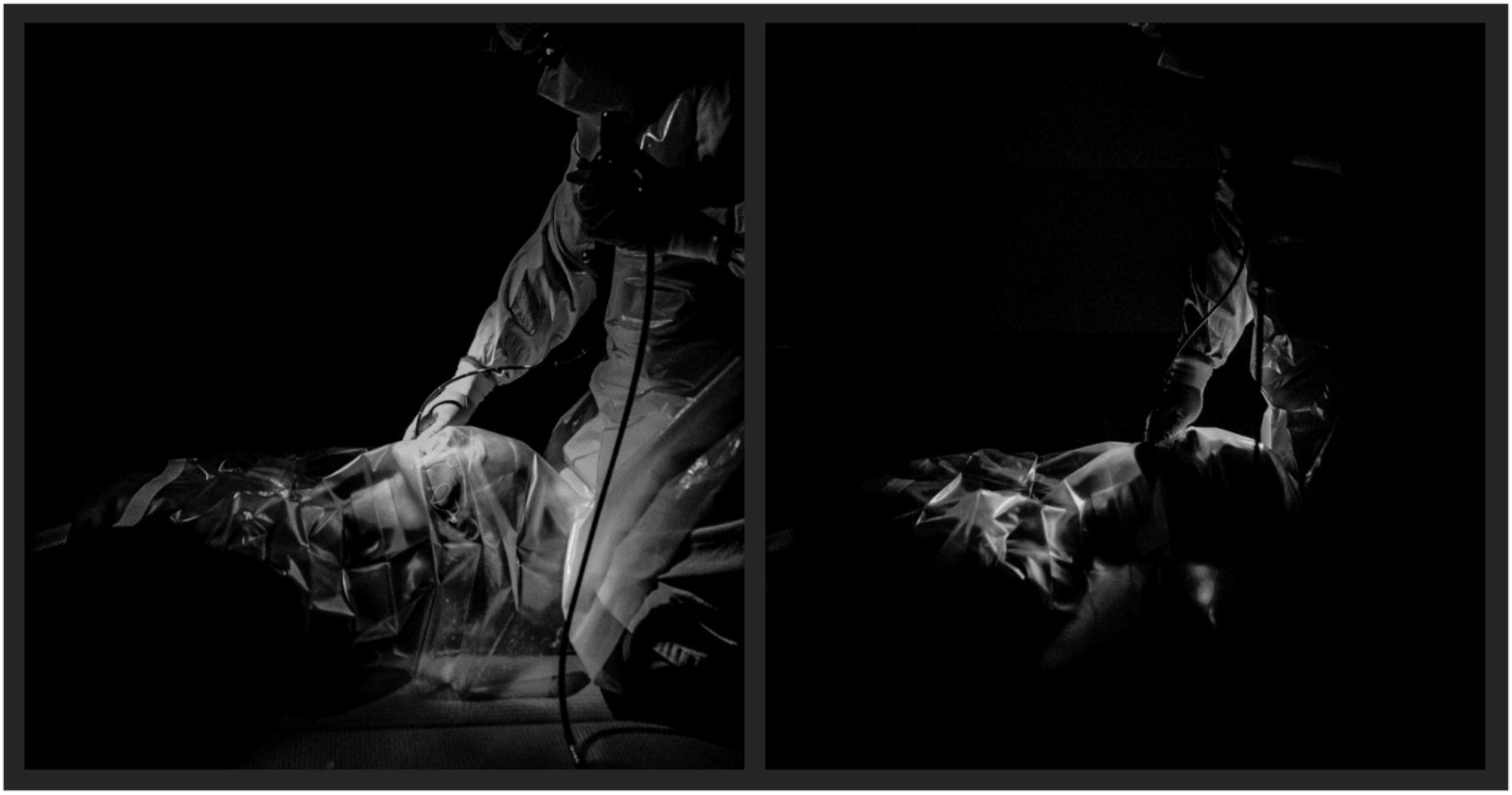
Protection from potential aerosol spread during upper endoscopy by using a covering drape.

During upper endoscopy using a covering clear drape the detector did not trace droplets or aerosol that were visualized by ultraviolet light.

## Discussion

SARS-CoV-2 is an unprecedented pandemic with high socio-economic consequences and burden for our health-care systems. The individual protection and safety precautions of our medical staff are crucial since HCW are at constant risk for infectious transmissions[12]. High-risk procedures like upper respiratory or gastrointestinal endoscopy are common in COVID-19 patients e.g. for diagnosis of COVID-19 associated pulmonary complications or upper gastrointestinal complication such as upper gastrointestinal hemorrhage or hepatobiliary interventions[19,20].

Besides viral transmission by droplets, there is growing evidence about aerosol transmission of SARS-CoV-2 which is dose dependent[6,14,21]. Whether invasive procedures like upper endoscopy generates aerosols in a crucial volume is a contentious issue of ongoing debates and has not been fully investigated[3,12]. Especially upper gastrointestinal endoscopy has been declared as an aerosol-generating investigation following recent research[22]. Besides that, experimental data support the possibility of transmission by aerosols even in the absence of AGPs[10]. Besides adequate PPE and increased distance to a patient (that is technical impossible for upper endoscopy), mechanical barriers may protect HCW during these invasive procedures[23].

In our study, we visualized both – potential aerosol and droplet spread during upper endoscopy during pharyngeal intubation. Our investigations indicate that a simple plastic drape placed over the patient has the potential to reduce the amount of aerosol and droplet spread and exposure of the investigator. This may be important for safety precautions since fenestrated drape cover for central venous access and plastic airway masks are widely available in emergency departments or intensive care units and its handling is routine. Moreover, this could be a comparatively cheap method of HCW protection particularly in low- or moderate-income countries. Previous research indicates that an oxygen mask or a surgical mask without a drape deflects aerosol spread and seems to be a comparatively less sufficient mechanical barrier than using a drape[18]. Performing endoscopy through a perforated drape and an oxygen mask for patient oxygenation appears practical in our investigations, not only since endoscopy through an oxygen mask alone is common in our practice.

In our opinion, this method of individual risk protection can be implemented in intensive care and resuscitation training sessions as safety trainings have been demonstrated to significantly reduce infection risks[1]. Besides that, invasive procedures should be performed by the most qualified investigators to reduce investigation-dependent contact time. Since the virus created a global public health threat, further investigation of patient and HCW safety is urgently needed.

## Data Availability

Included / on request.

## Literature

[1] Chou R, Dana T, Buckley DI, Selph S, Fu R, Totten AM. Epidemiology of and risk factors for coronavirus infection in health care workers: a living rapid review. Ann Intern Med 2020;173:120–36.

[2] Nolan JP, Monsieurs KG, Bossaert L, Böttiger BW, Greif R, Lott C, et al. European Resuscitation Council COVID-19 guidelines executive summary. Resuscitation 2020;153:45–55. https://doi.org/10.1016/j.resuscitation.2020.06.001.

[3] Chan VW-S, Ng HH-L, Rahman L, Tang A, Tang KP, Mok A, et al. Transmission of Severe Acute Respiratory Syndrome Coronavirus 1 and Severe Acute Respiratory Syndrome Coronavirus 2 During Aerosol-Generating Procedures in Critical Care: A Systematic Review and Meta-Analysis of Observational Studies. Crit Care Med 2021.

[4] Adams JG, Walls RM. Supporting the health care workforce during the COVID-19 global epidemic. Jama 2020;323:1439–40.

[5] Perlis RH. Exercising heart and head in managing coronavirus disease 2019 in Wuhan. JAMA Netw Open 2020;3:e204006–e204006.

[6] van Doremalen N, Bushmaker T, Morris DH, Holbrook MG, Gamble A, Williamson BN, et al. Aerosol and Surface Stability of SARS-CoV-2 as Compared with SARS-CoV-1. N Engl J Med 2020;382:1564–7. https://doi.org/10.1056/NEJMc2004973.

[7] Gralton J, Tovey E, McLaws M-L, Rawlinson WD. The role of particle size in aerosolised pathogen transmission: a review. J Infect 2011;62:1–13.

[8] Ong SWX, Tan YK, Chia PY, Lee TH, Ng OT, Wong MSY, et al. Air, surface environmental, and personal protective equipment contamination by severe acute respiratory syndrome coronavirus 2 (SARS-CoV-2) from a symptomatic patient. Jama 2020;323:1610–2.

[9] Klompas M, Baker M, Rhee C. What Is an Aerosol-Generating Procedure? JAMA Surg 2021;156:113–4.

[10] Klompas M, Baker MA, Rhee C. Airborne Transmission of SARS-CoV-2: Theoretical Considerations and Available Evidence. JAMA 2020;324:441–2. https://doi.org/10.1001/jama.2020.12458.

[11] Centers for Disease Control and Prevention. https://www.cdc.gov/coronavirus/2019-ncov/hcp/faq.html 2021, Accessed 6th March 2021 n.d. https://www.cdc.gov/coronavirus/2019-ncov/hcp/faq.html (accessed March 5, 2021).

[12] Tran K, Cimon K, Severn M, Pessoa-Silva CL, Conly J. Aerosol generating procedures and risk of transmission of acute respiratory infections to healthcare workers: a systematic review. PLoS One 2012;7:e35797–e35797. https://doi.org/10.1371/journal.pone.0035797.

[13] World Health Organisation. Modes of transmission of virus causing COVID-19: implications for IPC precaution recommendations. 2020, Accessed 1st March 2021 n.d. https://www.who.int/news-room/commentaries/detail/modes-of-transmission-of-virus-causing-covid-19-implications-for-ipc-precaution-recommendations.

[14] Tang S, Mao Y, Jones RM, Tan Q, Ji JS, Li N, et al. Aerosol transmission of SARS-CoV-2? Evidence, prevention and control. Environ Int 2020;144:106039. https://doi.org/10.1016/j.envint.2020.106039.

[15] Tran K, Cimon K, Severn M, Pessoa-Silva CL, Conly J. Aerosol-generating procedures and risk of transmission of acute respiratory infections: a systematic review. CADTH Technol Overv 2013;3.

[16] Hugonnet S, Pittet D. Transmission of severe acute respiratory syndrome in critical care: do we need a change? 2004.

[17] Gralnek IM, Hassan C, Beilenhoff U, Antonelli G, Ebigbo A, Pellisè M, et al. ESGE and ESGENA Position Statement on gastrointestinal endoscopy and the COVID-19 pandemic. Endoscopy 2020;52:483.

[18] Ott M, Milazzo A, Liebau S, Jaki C, Schilling T, Krohn A, et al. Exploration of strategies to reduce aerosol-spread during chest compressions: A simulation and cadaver model. Resuscitation 2020;152:192–8. https://doi.org/10.1016/j.resuscitation.2020.05.012.

[19] Koehler P, Cornely OA, Böttiger BW, Dusse F, Eichenauer DA, Fuchs F, et al. COVID-19 associated pulmonary aspergillosis. Mycoses 2020;63:528–34.

[20] Verweij PE, Gangneux J-P, Bassetti M, Brüggemann RJM, Cornely OA, Koehler P, et al. Diagnosing COVID-19-associated pulmonary aspergillosis. The Lancet Microbe 2020;1:e53–5.

[21] Banik RK, Ulrich A. Evidence of Short-Range Aerosol Transmission of SARS-CoV-2 and Call for Universal Airborne Precautions for Anesthesiologists During the COVID-19 Pandemic. Anesth Analg 2020;131.

[22] Sagami R, Nishikiori H, Sato T, Tsuji H, Ono M, Togo K, et al. Aerosols produced by upper gastrointestinal endoscopy: a quantitative evaluation. Off J Am Coll Gastroenterol ACG 2021;116:202–5.

[23] Gao Y, Xie J, Ye L-S, Du J, Zhang Q-Y, Hu B. Negative-Pressure Isolation Mask for Endoscopic Examination During the Coronavirus Disease 2019 Pandemic: A Randomized Controlled Trial. Clin Transl Gastroenterol 2021;12:e00314.

